# Effects of a diverse prebiotic fibre supplement on HbA1c, insulin sensitivity, and inflammatory biomarkers in pre-diabetes: a randomised placebo-controlled clinical trial

**DOI:** 10.1101/2024.01.09.24301052

**Authors:** C.V. Hall, J.L. Twelves, M. Saxena, L Scapozza, T. Gurry

**Affiliations:** Myota GmbH, London, UK; Lindus Health Limited, London, UK; William Harvey Research Institute, Barts NIHR Biomedical Research Centre, Queen Mary University of London, London, UK; Pharmaceutical Biochemistry Group, School of Pharmaceutical Sciences, University of Geneva, Switzerland

**Keywords:** pre-diabetes, gut microbiome, prebiotic fibre supplement, randomised controlled trial, placebo-controlled

## Abstract

Prebiotic fibre represents a promising and efficacious treatment to manage pre-diabetes, acting via complementary pathways involving the gut microbiome and viscosity-related properties. In this study, we evaluated the effect of using a diverse prebiotic fibre supplement on glycaemic, lipid, and inflammatory biomarkers in patients with pre-diabetes. Sixty-six patients diagnosed with pre-diabetes (yet not receiving glucose-lowering medications) were randomised into treatment (n = 33) and placebo (n = 33) interventions. Participants in the treatment arm consumed 20g per day of a diverse prebiotic fibre supplement and participants in the placebo arm consumed 2g per day of cellulose for 24 weeks. A total of 51 and 48 participants completed the week 16 and week 24 visits, respectively. The intervention was well-tolerated, with a high average adherence rate across groups. Our results extend upon previous work, showing a significant change in glycated haemoglobin (HbA1c) in the treatment group, but only in participants with lower baseline HbA1c levels (<6% HbA1c). Within the whole cohort, we showed significant improvements in insulin sensitivity, fasting plasma insulin, and C-reactive protein in the treatment group compared with the placebo. We did not observe any between-group differences in lipid profiles or other inflammatory cytokines. Together, our results show support for the beneficial effects of a diverse prebiotic fibre supplement on physiologically relevant biomarkers in pre-diabetes.

## Introduction

Pre-diabetes is an intermediate phase preceding type 2 diabetes mellitus (T2DM), where glucose tolerance, fasting glucose, and/or glycated haemoglobin (HbA1c) are impaired, but not yet within the diabetic range (1). Research efforts to date have primarily focused on interventions targeting patients with T2DM, owing to the growing burden on primary healthcare resources, and associated comorbidities including cardiovascular disease, kidney diseases, ocular dysfunction, and mental illness (2). However, evidence suggests that initiating treatment in pre-diabetes, where patients are earlier and less advanced in the diabetic disease pathway, may result in faster clinical improvements that are maintained long-term (3-5). Pre-diabetes therefore represents a critical period for early intervention, reducing the risk of developing T2DM and associated healthcare costs (6). While lifestyle forms a key part of this, diet and exercise interventions that target pre-diabetes are currently resource-intensive, challenging to maintain over time, and fail to address significant barriers to behaviour change experienced by those with pre-diabetes (7). There is therefore a pressing need to develop novel, scalable, cost-effective, and targeted interventions.

Emerging evidence suggests that the gut microbiome and its metabolites represent a promising target for pre-diabetes interventions, playing a key role in modulating glycaemic, metabolic, and inflammatory biomarkers (8). One of the key ways in which the gut microbiome is thought to exert these benefits is via microbial fermentation of dietary fibre in the colon, which typically results in the production of the Short Chain Fatty Acids (SCFAs) (9). SCFAs, including butyrate, propionate, and acetate, influence host health by maintaining intestinal barrier integrity (10, 11), modulating inflammation (12), and altering gene expression patterns in host tissue (13-15). In this manner, SCFAs can regulate the expression of pro- and anti-inflammatory cytokines, and influence glucose homeostasis and insulin sensitivity (16, 17). As an important modulator of SCFA production, recent work has studied the effects of prebiotic fibres on glycaemic status, insulin sensitivity, and inflammatory biomarkers in pre-diabetes. Many of these interventions have focused on single-ingredient supplements including resistant starch, resistant dextrin, fructooligosaccharides (FOS), and oligofructose-enriched inulin (18-20). While results are promising, it is clear that there are vastly different responses to interventions, with considerable individual variability (21). A possible source of underlying variability is via differences in microbial fibre fermentation capabilities (22). In this sense, the degree of efficacy from a prebiotic fibre supplement is related to how well it will be fermented by specific microbiota to produce SCFAs.

To overcome these constraints, work by us (22) and others (9) have shown that exposure to a broad range of prebiotic fibres can compensate for inter-individual variability in microbial fermentation abilities. A diversified fibre supplement could therefore lead to improved clinical outcomes. Independent to microbiome-mediated effects, dietary fibres with viscous physicochemical properties (e.g., arabinoxylan and beta-glucan) are well known for their role in reducing acute postprandial glucose and insulin responses (23). Interventions that leverage both prebiotic and postprandial glycaemic benefits represent a dual-pronged approach, acting via non-overlapping yet complementary mechanisms of action. In this study, we evaluated the effect of using a diverse prebiotic fibre supplement on metabolic, lipid, and inflammatory biomarkers in patients with pre-diabetes.

## Methods

### Study Design

This study was a parallel arm, individually randomised, single-blinded, placebo-controlled clinical trial with 1:1 treatment allocation ratio. The study was approved by Berkshire B NHS Human Research Ethics Committee in the United Kingdom (reference: 22/SC/0363). The study was registered on ClinicalTrials.gov (registration number: NCT05593926). Written informed consent was obtained from all participants in accordance with the Declaration of Helsinki.

### Study population

Eligible participants were men and postmenopausal women with a primary diagnosis of pre-diabetes, but not receiving treatment for type 2 diabetes, with baseline HBA1c within range of 5.8% (40mmol/mol) to 6.5% (48 mmol/mol), who were willing to complete study requirements, and had access to a smartphone or computer. Main exclusion criteria were participants receiving medication to treat type 1 or type 2 diabetes in the previous 6 months and current participation in a weight loss program or planned in the next 16 weeks. Detailed eligibility criteria are available in **Supplementary Note 1**.

### Study Intervention

Eligible participants who met the eligibility criteria were randomised using a web-based system (*Sealed Envelope*) in a ratio of 1:1 to either the treatment arm of 20g per day of a diverse fibre supplement (**Supplementary Note 2 & Supplementary Table 1**) or the placebo arm of 2g per day of cellulose, both unflavoured powders. The supplements were provided to the participants in single serve sachets and were taken at any time of the day with water or mixed into food or drink of their choice (e.g., breakfast cereal, tea, coffee). To monitor adherence, participants received a daily text message, asking them to confirm that they had taken the supplement. If they failed to consume the supplement on any given day, they were prompted to provide a reason. Participants were advised to consume the intervention every day for 16 weeks. While our primary endpoint was assessed at week 16, we also included an exploratory endpoint at week 24. Participants were advised to continue taking the intervention daily until their final study visit at week 24.

### Study Assessments

Participants attended clinic appointments at baseline, week 16 and week 24 after 10-12 hours of overnight fasting. They had blood pressure measurements, a fasting blood test, and an Oral Glucose Tolerance Test for Insulin Sensitivity (ISI-OGTT) performed at baseline, week 16, and week 24. For the ISI-OGTT (baseline and week 16 only), blood samples were taken at -15, 0, 30, 60, 90, and 120 min for the measurement of plasma glucose and insulin concentrations. From blood samples, cytokines were measured by using a cytokine multiplexing assay from Meso Scale Discovery. This assay measures IL-6, IL-8, IL-10 and TNF-*a* together in the same well of a plate. The full lipid profile, High Sensitivity CRP (CRP), plasma glucose, and HBA1c were all measured on an automated chemistry analyser (Siemens Advia 1800). Plasma insulin was measured on an automated immunoassay analyser (Siemens Advia Centaur XP). The insulin sensitivity index from the ISI-OGTT was calculated according to the equation derived by Matsuda & De Fronzo (24) in which insulin sensitivity is estimated by dividing a constant (10,000) by the square root of the product of fasting glucose (FBG) times, fasting insulin (FBI) times, mean glucose (MG) times and mean insulin (MI). ISI-OGTT = 10.000/(FPG*FPI)(MG*MI). The index is strongly correlated with insulin sensitivity derived using the euglycaemic clamp technique (24).

### Primary and secondary endpoints

The primary endpoint was the change in HbA1c from baseline to week 16. Key secondary endpoints included changes in HbA1c from baseline to 24 weeks, insulin sensitivity (measured using ISI-OGTT) from baseline to week 16, and changes in fasting plasma insulin (FPI), blood lipids (cholesterol [total, LDL, HDL], triglycerides), inflammatory biomarkers (Interleukin-6 [L-6], IL-8, IL-10, CRP, Tumor Necrosis Factor [TNF]-*a*), and diastolic and systolic blood pressure from baseline to 16 and 24 weeks.

### Tolerability and safety

Participants were asked to record whether they experienced any side-effects or medical events since starting the intervention, recorded via online surveys at the end of weeks 1-4, and on a monthly basis for the remainder of the trial. If they answered yes, adverse events were documented, reported, and reviewed for relatedness and expectedness within 24 hours. Discontinuations were captured and presented in **Fig 1**.

**Fig 1.**
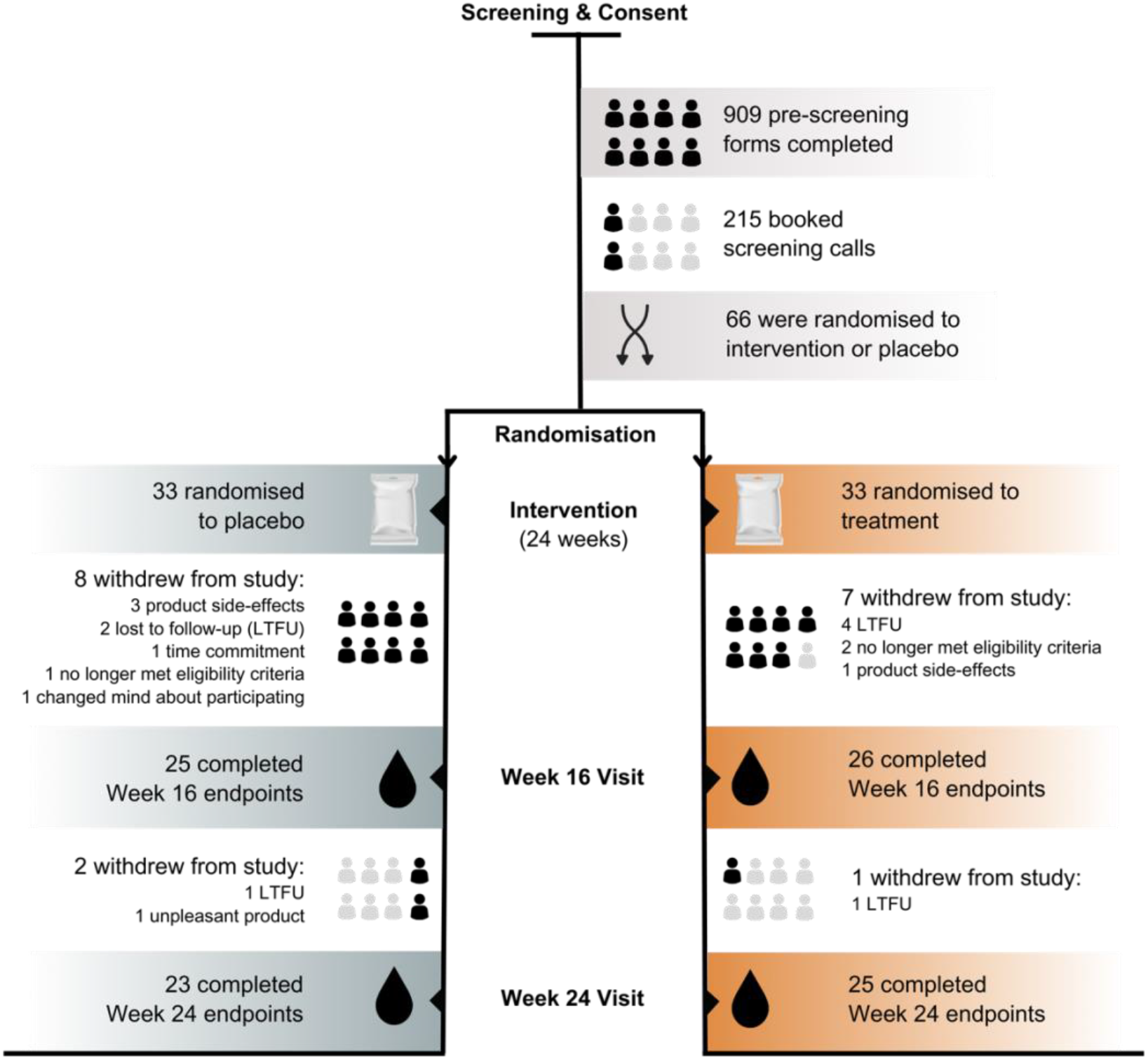
Clinical trial flow.

### Statistical analysis

Data were analysed using R software. Statistical analyses were performed based on an intention-to-treat protocol. For the primary endpoint (HbA1c) and ISI-OGTT, an unpaired t-test was used to compare the change from baseline to 16 weeks between the treatment and placebo. Two-way repeated measures Analysis of Variance (ANOVA) with one within-subject factor (time) and one between-subject factor (randomisation) were used to compare changes in secondary endpoints from baseline to week 16, baseline to week 24, and week 16 to 24. Post-hoc paired t-tests (Bonferroni corrected) were performed on significant main or interaction effects to compare differences between and within groups over time. For non-normally distributed outcome variables, robust two-way ANOVA with trimmed means were used, and Wilcoxon signed-rank tests were used for non-parametric post-hoc tests. Subgroup analyses included the effects of baseline HbA1c levels on the primary endpoint. Effects with p-values < 0.05 were considered statistically significant. As a pilot study, our sample size was informed from previous studies assessing fibre-based interventions in pre-diabetes cohorts. Results from previous studies (9, 18, 25) suggest that 60 participants is an adequate sample size to capture meaningful differences in our primary and secondary outcome measures. A sample size of 33 in each arm, accounting for an attrition rate of 10%, results in a power of 80% to detect effects of at least *d* = 0.74.

## Results

### Baseline patient characteristics

The study was conducted between 17 November 2022 to 9 October 2023. In total, 215 participants were screened, and 66 participants were randomised (33 treatment; 33 placebo) (**Fig 1**). The baseline characteristics were not significantly different across groups, with the exception of fasting plasma insulin (FPI) and triglycerides, which were not matched variables for inclusion or randomisation (**Table 1**). A total of 51 and 48 participants completed the week 16 and week 24 visits, respectively. The various reasons for participant withdrawals are presented in **Fig 1**.

**Table 1.** Baseline demographic and clinical characteristics.

|  | Treatment (n = 33) | Placebo (n = 33) | p-value |
| --- | --- | --- | --- |
| Sex (n female, (%)) | 10 (30%) | 8 (24%) | 0.78<sup>a</sup><br>($X^2 = 0.08$) |
| Age (years) | 55.7 ± 8.15 | 56.9 ± 8.15 | 0.62<sup>b</sup> |
| BMI (kg/m<sup>2</sup>) | 31.4 ± 4.18 | 30.8 ± 4.14 | 0.62<sup>b</sup> |
| HbA1c (%) | 6.12 ± 0.24 | 6.02 ± 0.19 | 0.09<sup>b</sup> |
| Fasting plasma glucose (mmol/L) | 5.72 ± 0.97 | 5.62 ± 0.69 | 0.66<sup>b</sup> |
| Fasting plasma insulin (μU/mL) | 17.6 ± 8.58 | 12.6 ± 7.18 | **0.03<sup>c</sup>** |
| Total cholesterol (mmol/L) | 5.07 ± 1.18 | 4.78 ± 1.06 | 0.36<sup>b</sup> |
| Triglycerides (mmol/L) | 1.61 ± 0.83 | 1.06 ± 0.44 | **0.004<sup>b</sup>** |
| LDL-C (mmol/L) | 2.77 ± 0.92 | 2.57 ± 0.79 | 0.38<sup>b</sup> |
| HDL-C (mmol/L) | 1.16 ± 0.33 | 1.16 ± 0.35 | 0.99<sup>b</sup> |
| IL-6 (ng/L) | 1.19 (0.47 - 5.28) | 1.25 (0.29-119.0) | 0.72<sup>c</sup> |
| IL-8 (ng/L) | 12.00 (7.60 - 91.2) | 14.00 (7.65 - 40.20) | 0.31<sup>c</sup> |
| IL-10 (ng/L) | 0.47 (0.18 - 1.36) | 0.34 (0.19 - 5.08) | 0.31<sup>c</sup> |
| CRP (mg/L) | 2.31 (0.06 - 28.00) | 1.51 (0.02 - 11.80) | 0.58<sup>c</sup> |
| TNF-α (ng/L) | 1.38 ± 0.25 | 1.41 ± 0.40 | 0.71<sup>b</sup> |
| Systolic blood pressure (mmHg) | 128 ± 15.9 | 130 ± 14.4 | 0.64<sup>b</sup> |
| Diastolic blood pressure (mmHg) | 81.9 ± 9.09 | 81.7 ± 10.1 | 0.95<sup>b</sup> |

Mean ± SD; Median (range); <sup>a</sup>Pearson's Chi-squared test; <sup>b</sup>Parametric t-test; <sup>c</sup>Wilcoxon signed-rank test.
BMI, body mass index; HbA1c, glycated haemoglobin; LDL-C, low density lipoprotein – cholesterol; HDL-C, high density lipoprotein – cholesterol; IL, interleukin; CRP, C-reactive protein; TNF- $\alpha$ , Tumor Necrosis Factor – alpha.

### Tolerability and safety

The intervention was well tolerated, with only two participants in the treatment group withdrawing from the study due to disliking the taste of the product. An additional three participants in the placebo group withdrew from the study due to product-related adverse events or not tolerating the product. There were no severe adverse events related to the treatment or placebo interventions. Adverse events reported were consistent with those typically seen in fibre-based interventions, including bloating, loose stools, and abdominal pain (**Supplementary Table 2**).

### Adherence

There was no statistical difference in adherence between the treatment and placebo arms (*F*_1,49_ = 1.19, *P* = 0.28). From baseline to week 16, average adherence was 89.3 ± 2.54% for the treatment group and 82.9 ± 3.79% for the placebo (mean ± SD). However, from week 16 to the exploratory endpoint at week 24, we observed a significant decrease in adherence over time in both the treatment (−8.33 ± 7.83%; *t*_26_ = 2.70, *P* = 0.01) and placebo groups (−6.79 ± 6.82%; *t*_26_ = 2.24, *P* = 0.03) (**Fig. 2a**).

**Fig 2.**
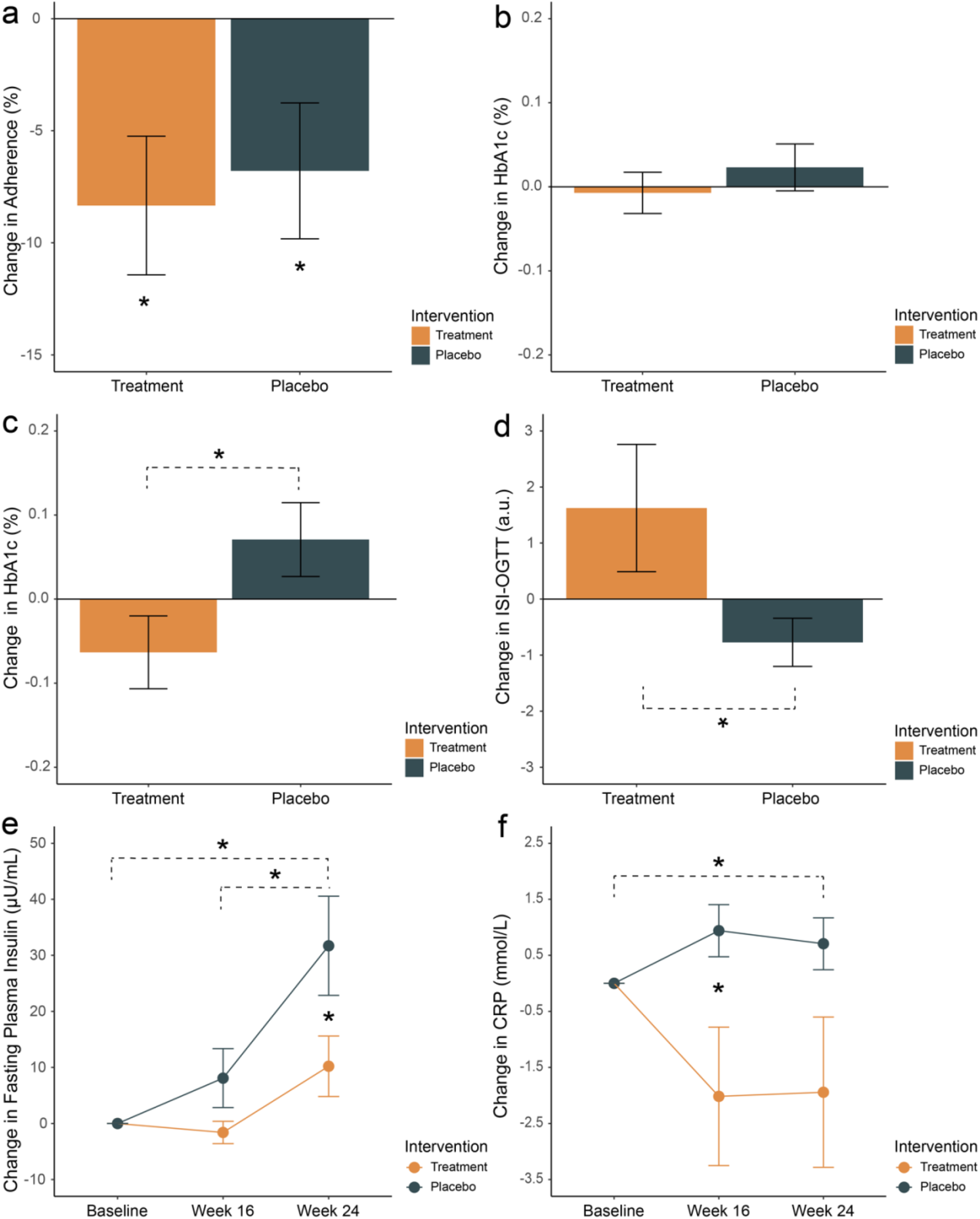
Change from baseline to week 16 in (a) adherence (%); (b) Glycated haemoglobin (HbA1c) (%); (c) HbA1c in a sub-group of participants with baseline HbA1c levels < 6.0%; and (d) insulin sensitivity (ISI-OGTT) (a.u.) in the treatment (orange) and placebo (green) groups. (e) Change from baseline to week 16 and 24 for (b) fasting plasma insulin (μU/ml) and (c) C-reactive protein (CRP) mmol/L. Mean and standard errors are shown. * indicates *p*-value < 0.05.

### Primary endpoint

The change in HbA1c from baseline to week 16 was not significantly different between the two groups (*P* = 0.37; treatment -0.02 ± 0.27% *vs*. placebo +0.04 ± 0.25%, mean ± SD) (**Fig. 2b**). In participants with a baseline HbA1c < 6.0% (i.e. earlier in the diabetic process), there was a significant reduction in HbA1c in the treatment group compared to the placebo (*P* = 0.05; treatment -0.17 ± 0.27 *vs*. placebo 0.07 ± 0.29, mean ± SD) (**Fig. 2c**).

### Secondary endpoints

#### Glycaemic outcomes

At week 16, insulin sensitivity, assessed via the ISI-OGTT, was significantly higher in the treatment group compared to the placebo (*P* = 0.03; treatment 1.62 ± 5.79 *v*. placebo -0.77 ± 2.11, mean ± SD) (**Fig. 2d**). For FPI, results of a two-way repeated measures ANOVA showed a significant main effect of group (*F*_1,46_ = 5.12, *P* = 0.03) and time (*F*_2,92_ = 13.31, *P* < 0.001) (**Fig. 2e**). In the placebo group, post-hoc paired t-tests (Bonferroni-corrected) showed a significant change from baseline to week 24 (*t*_22_ = -3.51, *P*_*FWE*_ = 0.01), and week 16 to week 24 (*t*_22_ = -2.94, *P* = 0.04). At week 24, the change in FPI in the placebo group was significantly higher compared to the treatment (*t*_12.6_ = -2.02, *P*_*FWE*_ = 0.05). There were no significant pairwise differences in FPI between baseline, week 16, or week 24 for the treatment group.

#### Inflammatory cytokines

For CRP, results of a robust two-way repeated measures ANOVA using trimmed means showed a significant main effect of group (*t* = 6.37, *P* = 0.02) and an interaction effect (*t* = 6.85, *P* = 0.05) (**Fig. 2f**). Pairwise post-hoc tests revealed that CRP was significantly lower at week 16 in the treatment group compared to placebo (*W* = 194, *P*_*FWE*_ = 0.03). In the placebo group, there was a significant change in CRP from baseline to week 24 (*W* = 168, *P*_*FWE*_ = 0.05). For TNF-*a*, there was a main effect of time (*F*_2,92_ = 7.83, *P* = 0.001), but no significant pairwise differences in the treatment or placebo groups. No differences were observed for IL-6, IL-8, or IL-10 (**Supplementary Fig. 1**).

#### Lipid profiles

Results from two-way repeated measures ANOVA in LDL-C showed a significant main effect of time (*F*_2,92_ = 32.8, *P* < 0.001) (**Supplementary Fig. 1**). For both the treatment and placebo groups, there was a significant increase in LDL-C from baseline to week 16 (treatment, *t*_22_ = -5.01, *P*_*FEW*_ = 0.0001; placebo, *t*_22_ = -3.11, *P*_*FWE*_ = 0.03), baseline to week 24 (treatment, *t*_22_ = -5.05, *P*_*FWE*_ = 0.0002; placebo, *t*_22_ = -4.54, *P*_*FWE*_ = 0.001), and week 16 to week 24 (treatment, *t*_22_ = -3.29, *P*_*FEW*_ = 0.02; placebo, *t*_22_ = -3.29, *P*_*FWE*_ = 0.02). Results from two-way repeated measures ANOVA in HDL-C showed a significant main effect of time (*F*_2,92_ = 7.54, *P* = 0.001) (**Supplementary Fig. 1**). Post-hoc tests showed no significant differences between baseline, week 16, or week 24 for the treatment or placebo groups. No significant differences were observed for total cholesterol, or triglycerides.

#### Blood pressure

No significant differences were observed for systolic or diastolic blood pressure across groups (**Supplementary Fig. 1**).

## Discussion

In this study, we showed a significant change in glucose metabolism and inflammatory responses in patients with pre-diabetes following a diverse prebiotic fibre intervention. Our primary outcome, the change in HbA1c from baseline to week 16, was not significantly different between groups (**Fig. 2b**). However, in participants with lower baseline HbA1c levels (< 6%), we observed significant improvements in the treatment group compared to the placebo (**Fig. 2c**). Consistent with these results, a recent study in pre-diabetes found that improvements in glycaemic outcomes in response to inulin supplementation could be predicted by better baseline glycaemic status (26). This has been supported in previous work, showing that supplementation with inulin improved outcomes in a pre-diabetes population with lower baseline HbA1c values (5.4 ± 0.1%, mean ± SD) (27) while another study using 4g oat beta-glucan found no effects on HbA1c or secondary outcomes in a pre-diabetes population with higher baseline values (HbA1c 6.4 ± 0.4%, mean ± SD) (28). These studies are consistent with our results, showing that patients who are earlier in the pre-diabetic disease pathway may be more likely to respond metabolically to prebiotic fibre interventions. One study presents a possible explanation for this, highlighting that pre-diabetes individuals with HbA1c levels marginally below the threshold for T2DM (between 6.4% - 6.5%) face the highest risk of comorbidities, specifically adverse cardiovascular events (29). In this study, authors propose that in the absence of a T2DM diagnosis, this population is less likely to receive timely medical intervention, despite their HbA1c status being only marginally better than that of individuals with T2DM (29). While there are fewer studies in pre-diabetes, there are a number of studies in T2DM that show significant improvements in HbA1c following prebiotic interventions (9, 21, 30, 31). However, the majority of patients in these studies were on stable anti-diabetic medication. The efficacy of prebiotic fibre interventions on HbA1c may therefore vary depending on the individual’s baseline glycaemic status, existing comorbidities, and medication use.

While HbA1c remains a cornerstone for diabetes management, it may not fully capture early metabolic disturbances in pre-diabetes which can sometimes precede significant changes in HbA1c (32). Our study suggests that secondary outcomes including insulin sensitivity, fasting insulin, and CRP may represent sensitive biomarkers to track responses to prebiotic fibre in a pre-diabetes population. Consistent with previous work, our data show that insulin sensitivity improves following prebiotic fibre intake (25, 33) (**Fig. 2d**), although a number of studies have not observed this trend (27, 34-36). We also observed a significant difference in fasting insulin at week 24 between groups, which appeared to be driven by increases over time in the placebo group (**Fig. 2e**). In the treatment group, increases in insulin sensitivity with unchanged fasting insulin levels may be attributed to improved insulin signalling pathways or increased responsiveness of insulin receptors in target tissues such as muscle, liver, and adipose tissue (33). Unchanged fasting insulin levels alongside increased insulin sensitivity may also indicate altered glucose dynamics, including a reduction in hepatic glucose output or increased glucose uptake by peripheral tissues (37).

Elevated levels of inflammatory cytokines are thought to be key contributors to beta cell impairment and insulin resistance (38, 39). Our results showed a significant difference in CRP at week 16 between groups, supporting a potential link between prebiotic fibre, inflammation, and metabolic health (**Fig. 2f**). CRP is recognised as an independent risk factor for cardiovascular disease (40, 41), and more recently has been linked to an increased risk of later development of diabetes (42, 43). The reduction in CRP in the treatment group is consistent with a meta-analysis of 14 RCTs showing a small (−0.37 mg/L) but significant reduction in circulating CRP after a dietary fibre or fibre-rich food intervention (44). In this current study, we observed a much larger reduction in CRP of -2.02 mg/L at week 16 (**Fig 2f**).

In our study, we used a diverse prebiotic fibre supplement which accounts for previously described inter-individual differences in microbial fermentation capabilities (22). While beyond the scope of this investigation, a possible mechanism involves prebiotic fibre acting on metabolic, and inflammatory outcomes via SCFA production. Locally, SCFAs play a role in maintaining intestinal barrier integrity by acting as the dominant energy source for colonocytes (10, 11), promoting mucous production (45), and regulating the secretion of interleukins (12), all of which reduce intestinal inflammation. SCFAs also act as histone deacetylase (HDAC) inhibitors and therefore alter gene expression patterns in host tissue (13-15). In this manner, SCFAs influence the expression of pro- and anti-inflammatory cytokines, which further regulates both local and systemic inflammation. This is particularly relevant in pre-diabetes where the control and regulation of inflammatory cytokines are likely to play a role in the advancement of insulin resistance and type 2 diabetes (38, 46, 47). Additionally, the SCFA butyrate promotes GLP-1 secretion in the colonic epithelium (via FFAR2/3 receptors) and intestinal gluconeogenesis, both of which may indirectly promote glucose homeostasis and insulin sensitivity (16, 17, 48). As this emerging field continues to develop, the importance of combining blood-based biomarkers with SCFA measurements will be critical to extend this work. Currently, meaningful quantification of SCFAs from patient samples remains a challenge, particularly in the context of clinical trials.

The strengths of our investigation include the rigorous placebo controlled RCT design, with high tolerability to the intervention, and no serious adverse events reported. However, several caveats need to be considered. As a pilot study, our sample size is relatively small. Future studies will require larger sample sizes to confirm our findings. Another consideration is that for the placebo, we used 2g of cellulose given that it is considered to have very little impact on microbial fermentation activities (49). However, adaptations of the gut microbiota over time have also been reported following cellulose ingestion, albeit with highly variable responses between individuals (50, 51). Future work should be carried out to understand the physicochemical properties of dietary fibres and their relationship to glucose homeostasis. As opposed to the more common choice of maltodextrin, the use of cellulose as a placebo could be considered a more conservative choice, potentially resulting in an underestimation of the quantitative impact of our prebiotic fibre supplement on HbA1c and insulin sensitivity. It is important to note that at our exploratory endpoint at week 24, we did not observe a continued improvement in HbA1c, in the lower baseline HbA1c sub-group or the full cohort. This result may, in part, be related to the fact that adherence decreased from weeks 16 to 24 in both groups (Fig. 2a). This result suggests that ongoing high adherence is required to ensure long-term efficacy. From a practical perspective, ensuring that the supplement can be easily integrated into a patient’s existing daily routine will be important to determine the extent of improvements experienced by prebiotic fibre interventions.

## Conclusion

Around 10-15% of pre-diabetics will progress to diabetes annually (6). Simple, safe, cost-effective, scalable, and targeted interventions are urgently needed to prevent those at higher risk progressing to T2DM. Here, we showed that a diverse prebiotic fibre supplement improved metabolic and inflammatory biomarkers over a 16-week intervention. Critically, our results showed that participants with lower baseline HbA1c levels experienced greater improvements in HbA1c. These results provide strong support that earlier stages of pre-diabetes offer a window of opportunity to effectively prevent the progression to more advanced diabetes. The broader improvements in insulin sensitivity, fasting insulin, and CRP across the whole cohort also indicate potentially beneficial changes that warrant further exploration. Larger and adequately powered clinical interventional studies will be necessary to confirm these findings and explore the broader changes beyond HbA1c that impact longer term disease progression and other health outcomes.

## Data Availability

Underlying data presented in the study are available upon reasonable request to the authors

## Author contributions

Conceptualization and methodology, C.V. Hall, J.L. Twelves, T. Gurry; Formal analysis: C.V. Hall and T. Gurry; Data curation: J.L. Twelves; Resources, T. Gurry; Writing—original draft, review, and editing, all authors; Funding acquisition, T. Gurry; Supervision, T. Gurry and M. Saxena.

