## Supplementary Material for "Effects of a diverse prebiotic fibre supplement on HbA1c, insulin sensitivity, and inflammatory biomarkers in pre-diabetes: a randomised placebo-controlled clinical trial"

**Hall et al.**

### Supplementary Notes

#### Supplementary Note 1. Participant eligibility criteria

Eligible participants were men and postmenopausal women with a primary diagnosis of pre-diabetes, but not receiving treatment for type 2 diabetes, with baseline HBA1c within range of 5.8% (40mmol/mol) to 6.5% (48 mmol/mol), who were willing to complete study requirements, and had access to a smartphone or computer. Exclusion criteria included participants receiving medication to treat type 1 or type 2 diabetes in the previous 6 months, a BMI >45 kg/m$^2$, loss of more than 5% body weight in the last 3 months, current participation in weight loss program or planned in the next 16 weeks, steroid use (except for over the counter NSAIDs, topical steroids, and inhalers), severe hepatic diseases, continuous antibiotic use for >3 days within 4 weeks prior to enrolment, continuous use of weight-loss drug within 3 months of study enrolment, gastrointestinal surgery (except for appendicitis or hernia surgery or co-existing pathology (Crohn's disease, coeliac disease, endometriosis, prostate cancer)), severe mental illness within 6 months prior to enrolment, receiving drug therapy to treat cholecystitis, peptic ulcers, urinary tract infection, acute pyelonephritis, urocystitis or hyperthyreosis, pituitary dysfunction, severe organic diseases including cancer, coronary heart disease, myocardial infarction or cerebral apoplexy, infectious diseases, including pulmonary tuberculosis and AIDS, history of alcoholism or substance misuse, significant dyslipidaemia, severe hypertension (>160/100 mmHg), use of any food supplements to control blood glucose (e.g., chromium picolinate) within two months of study enrolment. Eligible participants were identified through GP practices, and via social media campaigns. Participants were withdrawn from the trial if they started medications used to control their pre-diabetes.

#### Supplementary Note 2. Nutritional breakdown of diverse prebiotic fibre supplement

Each 20g daily sachet of supplement contains the following fibres:

- Wheat fibre (gluten free)
- Oat fibre (gluten free)
- Fructooligosaccharides
- Galactooligosaccharides
- Inulin
- Resistant dextrin
- Resistant maltodextrin
- Partially hydrolysed guar gum
- Guar gum

### Supplementary Tables

#### Supplementary Table 1. Nutritional composition of prebiotic fibre supplement

| Typical Values | Per 100g | Per portion (10g) | % DV |
|---|---|---|---|
| **Energy** | 185 kcal<br>774 kJ | 20 kcal<br>85 kJ | - |
| **Fat** | 0.0g | 0.0g | 0% |
| **Carbohydrate** | 100g | 10g | 3% |
| From sugar | 1.4g | 0.1g | 0.4% |
| Dietary fibre | 98.6g | 9.8g | 35% |
| **Protein** | 0.0g | 0.0g | 0% |
| **Salt** | 0.0g | 0.0g | 0% |

#### Supplementary Table 2. Adverse events

| Type of Adverse Event | *n* | Treatment (*n*) | Placebo (*n*) |
|---|---|---|---|
| Bloating/wind | 14 | 12 | 2 |
| Diarrhoea/loose stools | 10 | 9 | 1 |
| Abdominal pain/discomfort | 9 | 6 | 3 |
| HbA1c over 6.5% | 8 | 6 | 2 |
| Constipation | 7 | 3 | 4 |
| Appetite changes | 5 | 5 | 0 |
| Dizziness | 2 | 0 | 2 |
| Heartburn | 2 | 0 | 0 |
| High blood pressure | 1 | 1 | 0 |
| High temperature | 1 | 1 | 0 |
| Joint pain | 1 | 1 | 0 |
| Fatigue | 1 | 1 | 0 |
| Rise in cholesterol / glucose | 1 | 0 | 1 |
| Sensitive gums | 1 | 0 | 1 |

### Supplementary Figures

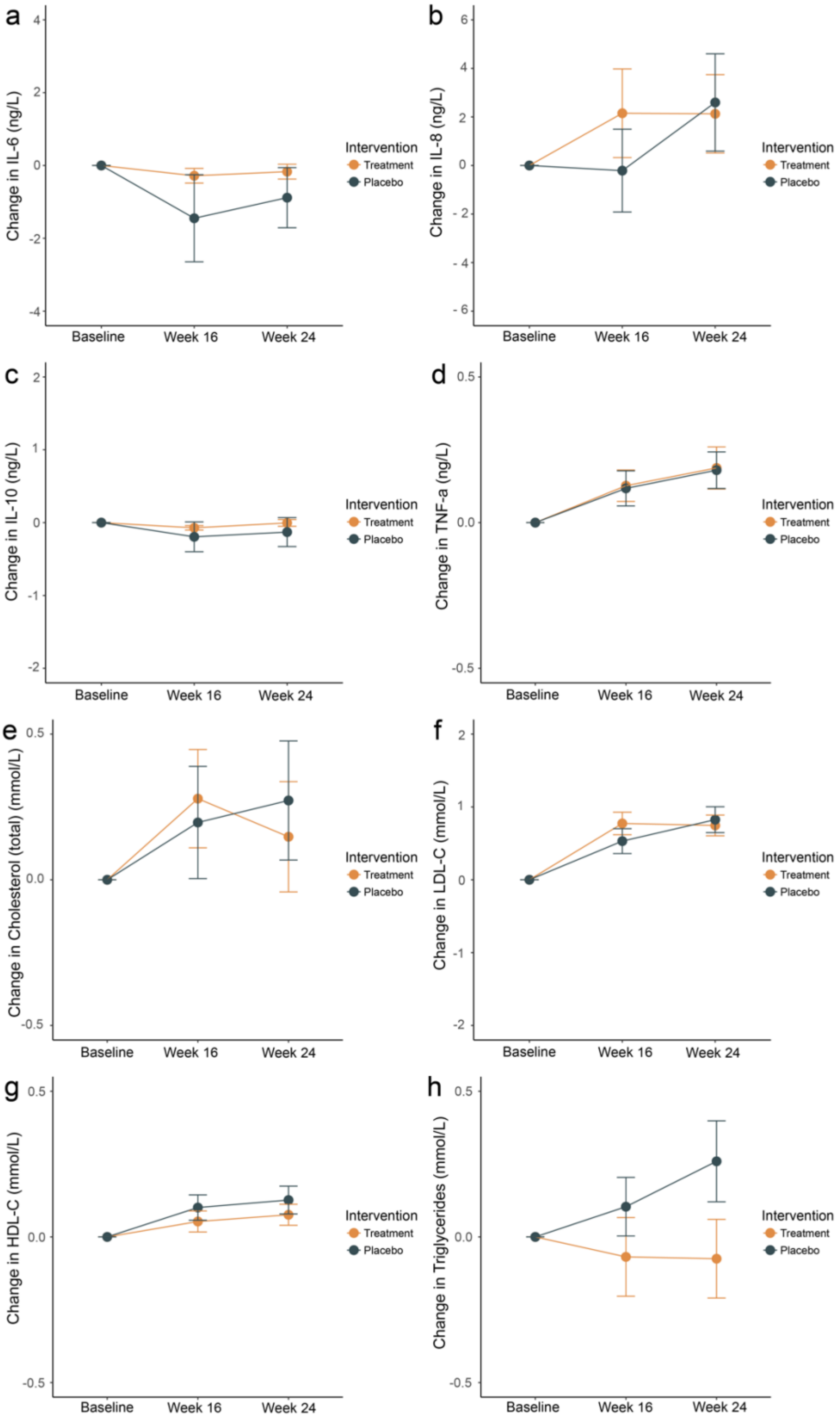


**Supplementary Fig 1.** Change from baseline to week 16 and 24 for (a) Interleukin (IL)-6, (b) IL-8, (c) IL-10, (d) TNF- $\alpha$, (e) cholesterol (total), (f) Low-density lipoprotein (LDL)-C, (g) High-density lipoprotein (HDL)-C, and (h) triglycerides for the treatment (orange) and placebo (green) groups. Mean and standard errors are shown.
